# Surgical strategy for atrial functional mitral regurgitation with atrial fibrillation

**DOI:** 10.1101/2024.09.23.24314258

**Authors:** Chunrong Bao, Ke Wei, Dongfang Zhao, Junwen Zhang, Ju Mei, Nan Ma

## Abstract

**Background:** With the growing prevalence of atrial fibrillation (AF), atrial functional mitral regurgitation (AFMR) combined with AF is expected to become a common clinical issue. We have summarized various surgical treatment strategies based on the degree of mitral regurgitation (MR) alongside rhythm control therapy for patients with AFMR and AF.

**Methods:** This retrospective study included 145 patients with AF and MR from January 2017 to January 2023. 33 patients with AF and moderate AFMR were designated as the moderate atrial regurgitation (MAR) group. 56 patients with AF and severe AFMR were designated as the severe atrial regurgitation (SAR) group. The remaining 56 patients with AF and severe primary MR were designated as the severe primary regurgitation (SPR) group. All patients in the MAR group underwent thoracoscopic AF procedure via a unilateral approach. Patients in the SAR and SPR groups underwent mitral valvuloplasty plus the Cox Maze IV procedure (CMP IV). Descriptive characteristics and outcomes were analysed.

**Results:** Twenty-three patients maintained sinus rhythm (SR) following thoracoscopic AF procedure in the MAR group at average 2.6±1.1 years follow-up. The degree of regurgitation improved in 26 patients, remained unchanged in 6 patients, and worsened in 1 patient. SR maintenance benefits MR reduction (P<0.0001) compared to the non-SR patients. There was no significant difference in the rate of SR maintenance following mitral valvuloplasty plus CMP IV between SAR (43 patients, 79.6%) and SPR (49 patients, 87.5%) groups. At the last follow-up, echocardiography in the SAR group showed 47 cases with no mitral regurgitation, 4 cases with mild regurgitation, and 1 case with moderate regurgitation. The left atrial diameter in the SAR group remained larger than in the SPR group (P<0.001).

**Conclusions:** Rhythm control therapy is the cornerstone treatment for AFMR patients with AF. Thoracoscopic AF procedure is effective and minimally invasive for moderate AFMR patients with AF. For severe AFMR patients with AF, we recommend CMP IV plus mitral valvuloplasty for safety and effectiveness.

## Introduction

Atrial functional mitral regurgitation (AFMR) is receiving increasing attention as a type of functional mitral regurgitation (MR) secondary to structural or functional abnormalities of the left atrium ^1^. Clinically, it is more common in patients with non-paroxysmal atrial fibrillation (AF) ^2,3^. Epidemiological studies have shown that the incidence of moderate to severe AFMR in patients with persistent AF is 4-8%, rising to 28% in those with AF lasting more than 10 years ^4,5^. With the growing prevalence of AF, AFMR combined with AF is expected to become a common clinical issue. However, there is still a lack of research (particularly surgical studies) addressing this combination. ^6,7^. Utilising a single treatment modality for MR with functional regurgitation is often unsatisfactory. Results from catheter ablation studies suggest that restoring sinus rhythm (SR) can reduce valve regurgitation through cardiac remodelling. However, the success rate of catheter ablation and the certainty of cardiac remodelling in reducing regurgitation remain concerning. We have summarized data from AFMR patients with AF treated at our centre, employing different treatment strategies based on the degree of MR alongside rhythm control therapy.

## Methods

### Patient Selection & Characteristics

This retrospective cohort study included 145 patients with AF and MR treated at the Department of Cardiothoracic Surgery, Xinhua Hospital Affiliated to Shanghai Jiao Tong University School of Medicine, from January 2017 to January 2023. Thirty-three patients with AF and moderate AFMR were designated as the moderate atrial regurgitation (MAR) group. 56 patients with AF and severe AFMR were designated as the severe atrial regurgitation (SAR) group. The remaining 56 patients with AF and severe primary MR were designated as the severe primary regurgitation (SPR) group. All patients received appropriate rate control, anti-hypertensives, and diuretic therapy before preoperative cardiac echo examination. The diagnostic criteria for AFMR are based on relevant literature: exclusion of organic mitral valve disease, left ventricular ejection fraction (LVEF) ≥50%, no abnormal left ventricular wall systolic activity, and left ventricular end-diastolic diameter <55 mm ^8,9^. The degree of mitral regurgitation was diagnosed according to guidelines and confirmed by two physicians.

The MAR group included 20 males and 13 females, with an average age of 70.27±6.46 years. Four cases had persistent AF and 29 had long-term persistent AF. The history of AF averaged 7.12±4.83 years, and the median CHA2DS2–VASc score was 3 points (0-6). Preoperative echocardiography showed moderate MR in 33 cases. The left atrium diameter was 48.15±4.19 mm, the left ventricular end-diastolic diameter was 49.91±3.29 mm, and LVEF was 56.39±4.90%. Preoperative cardiac function was NYHA grade I in 1 case, grade II in 17 cases, grade III in 12 cases, and grade IV in 3 cases. There were 22 patients with tricuspid regurgitation, 8 with coronary heart disease, 19 with hypertension, 10 with diabetes, and 3 with a history of stroke. Three patients had undergone catheter ablation. Baseline characteristics of MAR group are described in Table 1.

**Table 1.**
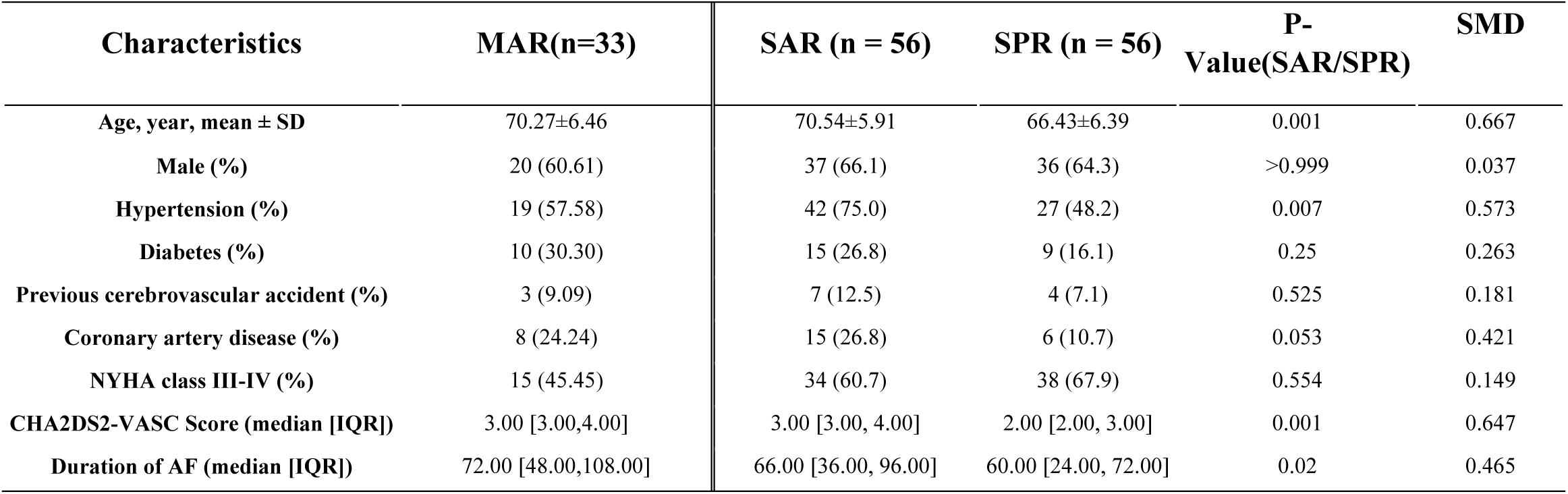
Patient Characteristics of three groups. . **Patients with moderate atrial functional mitral regurgitation (AFMR) and atrial fibrillation (AF) (MAR group). Patients with severe AFMR and AF (SAR group). Patients with severe primary mitral regurgitation and AF (SPR group).**

In the SAR group, there were 37 males and 19 females, with an average age of 70.54±5.91 years. All patients had long-term persistent AF with a history of 6.10±4.32 years. The median CHA2DS2–VASc score was 3 points (3-4). Thirty-eight patients had moderate or above tricuspid regurgitation, 15 had coronary heart disease, 42 had hypertension, 15 had diabetes, and 7 had a history of stroke. Four patients had a history of catheter ablation.

In the SPR group, there were 36 males and 20 females, with an average age of 66.43±6.39 years. Forty-four patients had long-term persistent AF, and 12 had persistent AF, with an AF history of 4.44±2.58 years. The median CHA2DS2–VASc score was 2 points (2-3). There were 23 patients with moderate or above tricuspid regurgitation, 6 with coronary heart disease, 27 with hypertension, 9 with diabetes, and 4 with a history of stroke. One patient had a history of catheter ablation. Baseline characteristics of both groups are described in Table 1.

### Surgical Procedures

All patients in the MAR group underwent thoracoscopic AF procedure via a unilateral approach as previously described [5, 6]. After anesthesia, patients were positioned in the right lateral decubitus position. Chest wall incisions were made at the 6th, 7th, 8th, or 6th, 8th, and 8th intercostal spaces near the inferior angle of the left scapula. The ablation routes included ablation of the left and right pulmonary veins, the posterior wall of the left atrium, the ablation line from the left pulmonary vein to the anterior mitral annulus (Dallas lesion), the ablation line from the left pulmonary vein to the resection edge of the left atrial appendage, and the ablation line between the two inferior pulmonary veins. The left atrial appendage was either resected with a stapler or clamped with an atrial appendage clip. Marshall’s ligament and epicardial autonomic ganglia were ablated by ablation pen. If AF could not be terminated after all steps, cardioversion was performed.

Patients in the SAR and SPR groups underwent mitral valvuloplasty plus the Cox Maze IV procedure (CMP IV). These procedures were performed under general anesthesia and extracorporeal circulation. A minimally invasive approach was used to establish extracorporeal circulation through the femoral artery and vein, with a small incision made in the fourth intercostal space on the right chest wall. All mitral annuloplasties were performed with a prosthetic ring. Additional techniques included artificial chordae, partial excision of the posterior leaflets, junctional suturing, and edge-to-edge suturing. If mitral annuloplasty failed, mitral valve replacement was performed. CMP IV included standard ablation routes for both the left and right atria, completed with a bipolar clamp, with isthmus ablation supplemented by cryoenergy. The left atrial appendage was resected or sutured in a double-layered linear fashion. Concomitant procedures included tricuspid valvuloplasty, coronary artery bypass grafting.

### Postoperative Care and Follow-up

Postoperatively, all patients received anticoagulation therapy with warfarin. If patient’s heart rate exceeded 70 beats/min, oral amiodarone would be administered for 3 months. Patients unable to maintain SR were treated with rate control. Recurrent AF was managed with cardioversion under adequate anticoagulation. Follow-up was conducted through outpatient visits and telephone calls, including symptom evaluation and 24-hour dynamic electrocardiograms at 3, 6, and 12 months, and every 6 months thereafter. Any documented atrial tachyarrhythmia episodes lasting 30 seconds or longer were considered a recurrence after a 3-month blanking period. Postoperative mitral valve function was assessed by echocardiography at 3, 6, and 12 months, and at least annually thereafter.

### Statistical Analysis

Statistical analysis was performed using R (version 4.3.2). Measurements are expressed as mean ± standard deviation and compared using the t-test. Count data are expressed as rates (percentages) and compared using Fisher’s exact test. Kaplan-Meier (KM) curve was utilized to analyze the success rates of AF treatments, while differences in success rates across groups were assessed using the log-rank test, with a P-value of less than 0.05 considered statistically significant. Left atrial diameter values over time are shown in box plots, with median, quartiles, and outliers. Differences were tested using repeated measures ANOVA. Changes in MR severity are displayed in stacked bar charts, indicating patient counts and percentages. Differences across stages were analyzed using Chi-square tests, with statistical significance set at P<0.05. Factors associated with recurrence following AF ablation were identified through an extensive review of the literature and previous experiences, and included in a multivariate regression model following univariate analysis where a P-value less than 0.1 was significant. A multivariate Cox proportional hazards regression model was employed to evaluate risk factors for AF recurrence, providing a comprehensive analysis of potential predictors.

### Ethical review

This study has been approved by the Medical Ethics Committee of Xinhua Hospital Affiliated to Shanghai Jiaotong University School of Medicine, and informed consent was obtained from all participants.

## Results

### Clinical Outcomes of AF Combined with Moderate AFMR

Thoracoscopic AF procedure via a unilateral approach was successfully performed in all patients in the MAR group. There was no conversion to thoracotomy and no perioperative mortality. No blood products were transfused, and no serious postoperative complications occurred. SR was maintained in 30 patients (90.9%) at discharge. Echocardiography before discharge showed no mitral regurgitation in 18 cases, mild mitral regurgitation in 10 cases, and moderate mitral regurgitation in 4 cases. The left atrium diameter (47.06±3.99 mm), left ventricular end-diastolic diameter (49.58±3.16 mm), and LVEF (55.73±5.40%) were not statistically different from pre-procedure measurements.

The last follow-up was in June 2023, with follow-up durations ranging from 1 to 6 years (average: 2.6±1.1 years). Twenty-three patients (69.70%) maintained SR, confirmed by 24-hour electrocardiography. The Kaplan-Meier curve illustrating freedom from AF recurrence is shown in Figure 1A. At the last follow-up, 17 patients had no mitral regurgitation, 9 had mild mitral regurgitation, 6 had moderate mitral regurgitation, and 1 had severe mitral regurgitation. The degree of regurgitation improved in 26 patients compared to pre-surgery levels, remained unchanged in 6 patients, and worsened in 1 patient. The left atrium diameter (46.42±4.40 mm), left ventricular end-diastolic diameter (49.94±3.32 mm), and LVEF (58.64±3.88%) were not statistically different from pre-procedure measurements.

**Figure 1.**
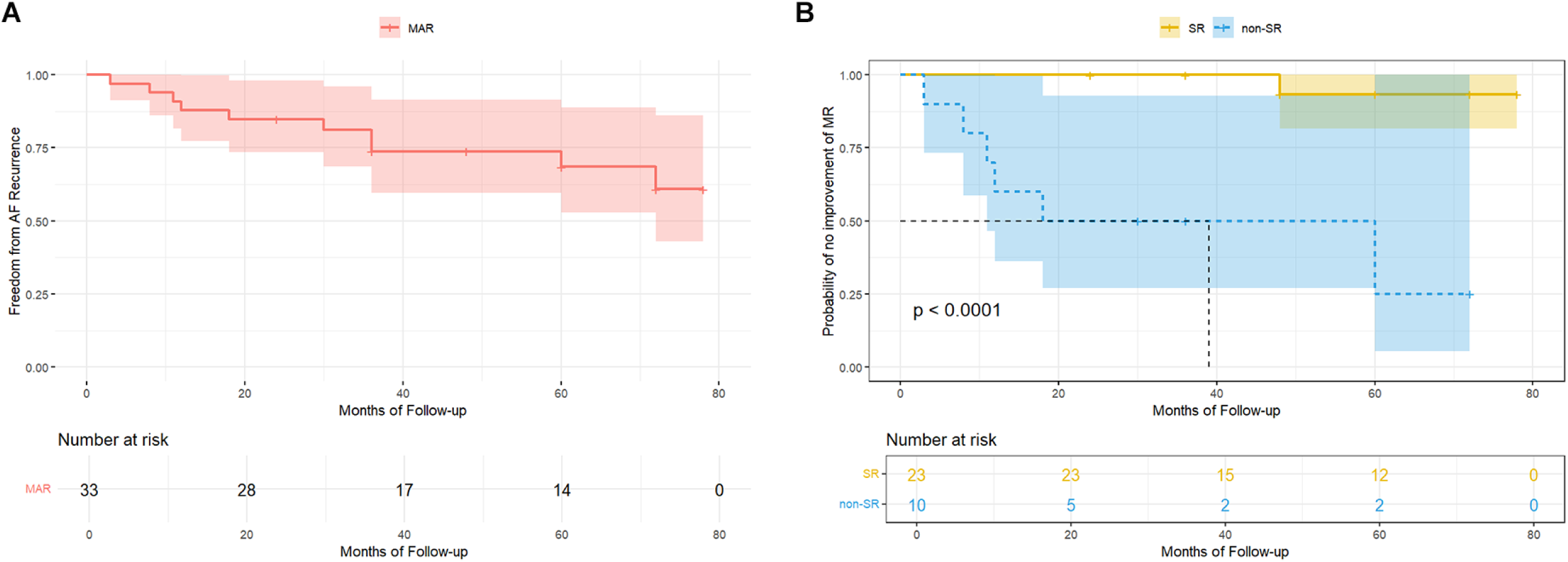
Six-year Kaplan-Meier curve for freedom from AF recurrence and comparison of mitral regurgitation reduction between SR and non-SR patients. 1A. Six-year Kaplan-Meier curve for atrial fibrillation (AF) patients with moderate atrial functional mitral regurgitation (AFMR) freedom from AF recurrence. 1B Sinus rhythm (SR) maintenance of AF patients with moderate AFMR benefits mitral regurgitation reduction (P<0.0001) compared to the non-SR patients.

### Effects of Rhythm Control on Cardiac Remodeling and Mitral Regurgitation in the MAR Group

Patients in the MAR group were divided into two cohorts based on their ability to maintain SR post-surgery. SR maintenance benefits MR reduction (P<0.0001) compared to the non-SR patients (Figure 1B). The left atrial diameters of the SR and non-SR patients were 47.26±4.06 mm vs. 50.20±3.94 mm before surgery (P=0.063) and 46.48±4.00 mm vs. 48.40±3.84 mm at discharge (P=0.209), showing no statistical difference between the groups. At the last follow-up, the left atrial diameter was 45.04±3.48 mm in the SR patients vs. 49.60±4.81 mm in the non-SR patients (P=0.004), a statistically significant difference (Figure 2A). The Corresponding changes in MR degree in SR and non-SR patients is shown in Figure 2B.

**Figure 2.**
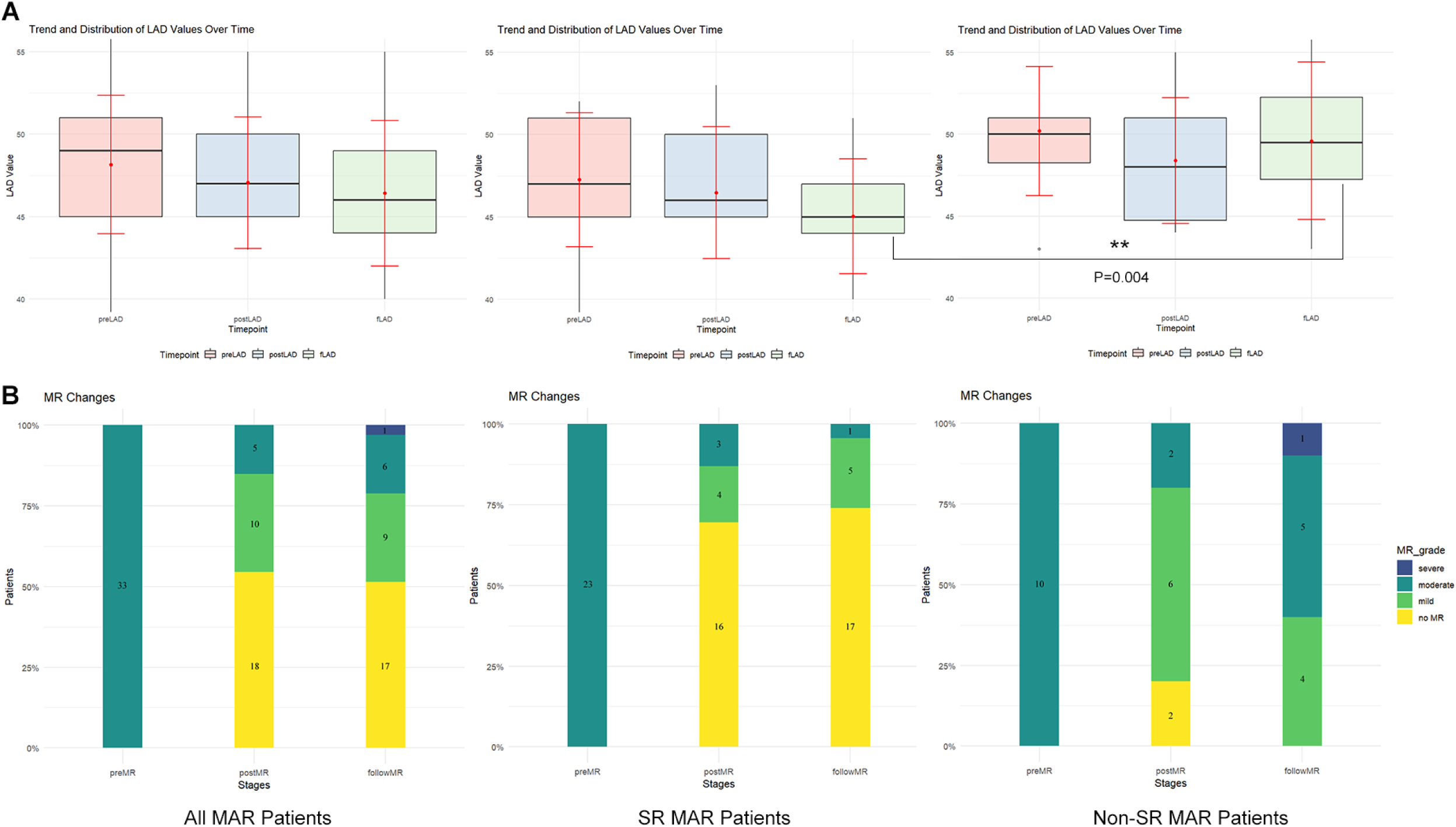
Comparison of left atrial diameters between SR patients and non-SR patients and the changes in the degree of mitral regurgitation following thoracoscopic AF procedure. 2A. Comparison of left atrial diameters between sinus rhythm (SR) patients and non-SR patients with moderate atrial functional mitral regurgitation (AFMR) and atrial fibrillation (AF) at different time points (pre-procedure, post-procedure, and at the last follow-up). At the last follow-up, the left atrial diameter was 45.04±3.48 mm in SR patients vs. 49.60±4.81 mm in non-SR patients (P=0.004), a statistically significant difference. 2B. The corresponding changes in the degree of mitral regurgitation in SR and non-SR patients with moderate AFMR and AF.

### Clinical Outcomes of AF Combined with Severe AFMR

All 56 patients in the SAR group completed the valve procedure, CMP IV, and other concomitant procedures. Two patients died within 30 days post-surgery due to postoperative ARDS and pulmonary infection. Postoperative complications included bleeding, low cardiac output, pulmonary infection, and acute kidney injury. Three cases (5%) required postoperative electrical cardioversion therapy. At discharge, 50 patients (92.6%) were in SR; 50 cases had no mitral regurgitation and 2 had mild mitral regurgitation. The left atrium diameter was 48.31±2.60 mm, significantly smaller than pre-surgery (P<0.001), while the left ventricular end-diastolic diameter (49.41±2.93 mm, P=0.056) and LVEF (54.87±5.46%, P=0.385) showed no significant changes from pre-surgery.

The mean follow-up period was 2.6±1.1 years. At the last follow-up, 43 patients (79.6%) in the SAR group maintained SR (KM curve shown in the Figure 3). Echocardiography showed no mitral regurgitation in 47 cases, mild regurgitation in 4 cases, and moderate regurgitation in 1 case. The left atrium diameter was 48.46±3.80 mm, significantly smaller than pre-surgery (P<0.001), while the left ventricular end-diastolic diameter (51.52±3.54 mm, P=0.225) and LVEF (54.80±4.93%, P=0.310) showed no significant differences from pre-surgery measurements.

**Figure 3.**
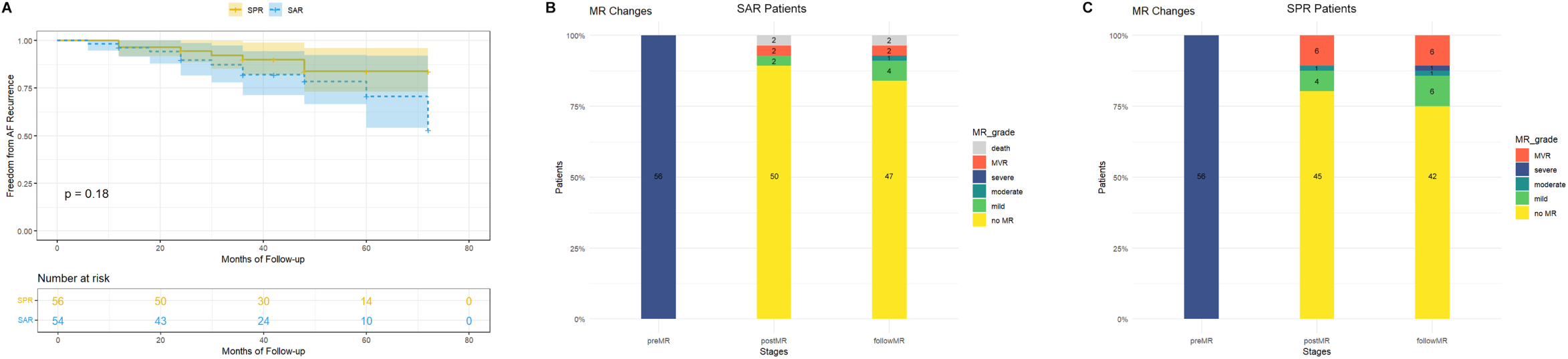
Kaplan-Meier (KM) curves of SR maintenance rate and the changes in the degree of mitral regurgitation following mitral valvuloplasty plus the Cox Maze IV procedure. 3A. KM curves of sinus rhythm (SR) maintenance rate for severe atrial functional mitral regurgitation patients with atrial fibrillation (AF) and severe primary mitral regurgitation patients with AF. The KM curves showed no statistical difference between the two groups following Cox Maze IV procedure. 3B. the degree of mitral regurgitation in atrial fibrillation (AF) patients with severe atrial functional mitral regurgitation and those with severe primary mitral regurgitation before the procedure.3C the degree of mitral regurgitation in AF patients with severe atrial functional mitral regurgitation and those with severe primary mitral regurgitation after the procedure.

### Comparison of Perioperative Data between SAR and SPR Groups

The SAR group patients were significantly older and had longer AF durations and more preoperative comorbidities than the SPR group. Detailed information is presented in Table 1. In the SAR group, 53 patients underwent mitral valvuloplasty with rings of varying sizes, including 11 patients with No. 28 ring, 24 patients with No. 30 ring, 16 patients with No. 32 ring, and 2 patients with No. 34 ring. Three patients underwent mitral valve replacement with bioprosthetic valves. Concomitant procedures included tricuspid valvuloplasty (35 cases), coronary artery bypass grafting (8 cases), and left ventricular outflow tract obstruction (2 cases). In the SPR group, 50 patients underwent mitral valvuloplasty with rings of varying sizes, including 10 patients with No. 28 ring, 23 patients with No. 30 ring, 15 patients with No. 32 ring, and 2 patients with No. 34 ring. Six patients underwent mitral valve replacement (3 bioprosthetic and 3 mechanical valves). Concomitant procedures included tricuspid valvuloplasty (19 cases) and coronary artery bypass grafting (2 cases). All patients in both groups completed CMP IV. There was no perioperative mortality in the SPR group. The SAR group had a slightly higher incidence of postoperative complications (Table 2). Fifty patients (92.6%) in the SAR group and 51 patients (91.1%) in the SPR group were in SR at discharge, with no significant difference between the groups (P=0.368). At the last follow-up, 43 patients (79.6%) in the SAR group and 49 patients (87.5%) in the SPR group maintained SR. The KM curves for the SR maintenance rate showed no statistical difference between the groups (Figure 3A).

**Table 2.** Postoperative complications of patients with severe atrial functional mitral regurgitation and atrial fibrillation (AF) (SAR group) and patients with severe primary mitral regurgitation and AF (SPR group).

| Postoperative Event | SAR (n = 56) | SPR (n = 56) | P.value |
| --- | --- | --- | --- |
| Stroke | 2 | 0 | 0.495 |
| Pneumonia | 6 | 3 | 0.489 |
| Poor wound healing | 1 | 0 | >0.999 |
| Prolonged ventilation | 7 | 4 | 0.527 |
| Acute renal failure | 4 | 1 | 0.364 |
| Blood product transfusion | 7 | 2 | 0.162 |
| Reoperation for bleeding | 1 | 0 | >0.999 |
| New pacemaker | 3 | 1 | 0.618 |
| Mortality within 30 days | 2 | 0 | 0.495 |
| Stays, day (median [IQR]) | 10.00 [9.00, 12.75] | 7.00 [7.00, 8.25] | <0.001 |

### Comparison of Mitral Regurgitation and Cardiac Remodeling Between SAR and SPR Groups

Preoperative echocardiography revealed that in the SAR group, 53 cases were Carpentier type I and 3 were type IIIb MR. In the SPR group, there were 5 cases of Carpentier type I and 51 cases of type II MR. There was no significant difference in left atrial diameter between the groups (P=0.532). The SPR group had a significantly larger left ventricular diameter (P<0.001) and lower ejection fraction (P<0.001) compared to the SAR group. Before discharge, 50 cases in the SAR group had no mitral regurgitation, and 2 had mild regurgitation. In the SPR group, 45 cases had no mitral regurgitation, 4 had mild regurgitation, and 1 had moderate regurgitation. The left atrial diameter in the SAR group was larger than in the SPR group (P<0.001). The ejection fraction in the SAR group was lower than in the SPR group, with no significant difference in left ventricular diameter (P=0.077). At the last follow-up, echocardiography showed that in the SAR group, 47 cases had no mitral regurgitation, 4 had mild regurgitation, and 1 had moderate regurgitation. In the SPR group, 42 cases had no mitral regurgitation, 6 had mild regurgitation, 1 had moderate regurgitation, and 1 had severe regurgitation. The left atrial diameter in the SAR group remained larger than in the SPR group (P<0.001), with no significant difference in left ventricular diameter (P=0.263) and ejection fraction (P=0.001). Detailed data are presented in Table 3. The changes in MR degree in the SAR and SPR groups are shown in Figure 3B and 3C.

**Table 3.** Comparison of cardiac remodeling between patients with severe atrial functional mitral regurgitation and atrial fibrillation (AF) (SAR group) and patients with severe primary mitral regurgitation and AF (SPR group). preLAD, pre-operative left atrial diameter; preLVEDD, pre-operative left ventricular end-diastolic diameter; preLVEF, pre-operative left ventricular ejection fraction; postLAD, post-operative left atrial diameter; postLVEDD, post-operative left ventricular end-diastolic diameter; preLVEF, post-operative left ventricular ejection fraction; fLAD, follow-up left atrial diameter; fLVEDD, follow-up left ventricular end-diastolic diameter; fLVEF, follow-up left ventricular ejection fraction

| Characteristics | SAR (n = 54) | SPR (n = 56) | P-Value | SMD |
| --- | --- | --- | --- | --- |
| preLAD, mm, mean ± SD | 54.61±5.57 | 53.95±5.22 | 0.519 | 0.123 |
| preLVEDD, mm, mean ± SD | 50.61±3.85 | 56.30±5.84 | <0.001 | 1.150 |
| preLVEF (%), mean ± SD | 55.65±3.77 | 51.48±4.85 | <0.001 | 0.960 |
| postLAD, mm, mean ± SD | 48.31±2.60 | 46.45±3.07 | 0.001 | 0.656 |
| postLVEDD, mm, mean ± SD | 49.41±2.93 | 50.64±4.18 | 0.077 | 0.342 |
| postLVEF (%), mean ± SD | 54.87±5.46 | 49.25±5.87 | <0.001 | 0.992 |
| fLAD, mm, mean ± SD | 48.46±3.80 | 46.02±2.94 | <0.001 | 0.720 |
| fLVEDD, mm, mean ± SD | 51.52±3.54 | 52.29±3.61 | 0.263 | 0.214 |
| fLVEF (%), mean ± SD | 54.80±4.93 | 51.27±5.94 | 0.001 | 0.646 |
| 498 preLAD, pre-operative left atrial diameter; preLVEDD, pre-operative left ventricular end-diastolic<br>499 diameter; preLVEF, pre-operative left ventricular ejection fraction; postLAD, post-operative left atrial<br>500 diameter; postLVEDD, post-operative left ventricular end-diastolic diameter; preLVEF, post-501 operative left ventricular ejection fraction; fLAD, follow-up left atrial diameter; fLVEDD, follow-up<br>502 left ventricular end-diastolic diameter; fLVEF, follow-up left ventricular ejection fraction |  |  |  |  |

### Analysis of Risk Factors for Recurrence of AF

Cox multivariate analysis indicated that AF recurrence in the MAR group was associated with a lack of regurgitation reduction (HR 0.0369; 95% CI 0.0026-0.5156; P=0.0142), reduction of left atrial diameter (HR 0.01865; 95% CI 0.0007-0.5016; P=0.0177), and was independent of age, CHA2DS2–VASc score, preoperative or postoperative left atrial diameter. In severe MR, the risk factors for AF recurrence in the SAR group included postoperative left atrial diameter greater than 50 mm (HR 4.081; 95% CI 1.109-15.016; P=0.0344), with no association with age, AF duration, CHA2DS2–VASc score, preoperative left atrial diameter, or residual mitral regurgitation. In the SPR group, AF recurrence was associated with a postoperative left atrial diameter greater than 50 mm (HR 12.480; 95% CI 1.288-120.898; P=0.0294) and residual mitral regurgitation (HR 16.358; 95% CI 1.604-166.850; P=0.0183), with no relation to age, AF duration, CHA2DS2–VASc score, or preoperative left atrial diameter. This study did not identify risk factors for the recurrence of mitral regurgitation in either group.

## Discussion

AF and MR are progressive conditions that exacerbate each other, leading to more severe symptoms, rapid disease progression, and serious consequences such as heart failure ^10^. Timely and effective treatment is particularly important when AF is combined with MR ^11^. We selected different treatment strategies based primarily on the degree of mitral regurgitation, which were all predicated on the treatment of AF. Our data suggests that for moderate AFMR combined with AF, rhythm control via thoracoscopic AF procedure yields satisfactory results. Maintenance of SR improves cardiac remodeling and reduces mitral regurgitation.

For severe AFMR combined with AF, we employed a combination of rhythm control and vavular intervention using the classic CMP IV surgical technique in conjunction with mitral valvuloplasty. This approach aligns with standard surgical guidelines for treating AF combined with severe MR. However, most previous evidence pertains to primary mitral regurgitation combined with AF, not AFMR combined with AF ^12,13^. Our study demonstrates that the safety and efficacy of CMP IV combined with mitral valvuloplasty for severe AFMR with AF are comparable to the outcomes in primary mitral regurgitation with AF. AF is the initiating factor of AFMR, making it the primary target for treatment. In cases of functional mitral regurgitation, merely treating the regurgitation without addressing the primary disease often fails to yield favourable outcomes ^14^. In cases of moderate regurgitation, the structural changes in the atrium and mitral valve are reversible ^15,16^. Although rhythm control may reduce or eliminate mitral regurgitation through cardiac remodeling, the efficiency remains unsatisfactory ^17,18^. The effectiveness of rhythm control is crucial in the management of AF. However, a large proportion of patients with AFMR have long-standing persistent AF, significant left atrium enlargement, high CHA2DS2–VASc scores, and other factors that impede successful treatment or maintenance of SR ^19^. Previous research has shown that thoracoscopic surgery may be more effective than catheter ablation in SR maintenance ^20^. The thoracoscopic technique provides high-quality pulmonary vein ablation, directly ablates the Marshall Ligament and autonomic ganglia from the epicardium, and enables left atrial appendage excision. Our modified procedure includes the isolation of the left atrial posterior wall to further improve success rates ^21^. Our data shows favourable results for AF patients with moderate AFMR, although subsequent catheter ablation may be required for patients with unsuccessful outcomes. This approach is reasonable for AFMR patients who require a more robust method to maintain SR and interrupt the cycle of AF, MR, and heart failure ^22^.

Patients with severe AFMR are more challenging to manage for clinicians. We therefore utilized CMP-IV, a classic surgical technique for treating AF, which has the highest single success rate in clinical practice. Despite their increased age, complexity, and potential complications, patients with severe AFMR still require treatment options that offer a high degree success in maintaining SR. As such, these patients would benefit the most from individualized ablation strategies or further minimally invasive procedures.

AFMR is mitral regurgitation secondary to structural or functional abnormalities of the left atrium, leading to mitral annulus enlargement and an imbalance between annulus and leaflet area. Although functional in nature, AFMR may still involve abnormal mitral valvular structure ^23,24^. Studies suggest that there is potential for mitral valve with structural abnormalities to recover with improved cardiac remodeling ^25^. However, for most patients with severe AFMR, surgical valvuloplasty is a neccesity due to underlying structural abnormalities ^26,27^. In patients with AFMR, even with successful AF treament, cardiac remodeling alone may not achieve the necessary changes in the left atrium and mitral valve. From the perspective of AF management, patients with AFMR and AF are often refractory to catheter ablation, thoracoscopic AF procedure, and even hybrid procedures. Therefore, treatment with CMP-IV should be prioritized, as it is more reasonable to undertake concomitant mitral valvuloplasty with its more definitive effect, over over cardiac remodeling with uncertain outcomes in MR ^28^. According to current guidelines, severe MR should be treated from a valvular management perspective. Without treatment, residual mitral regurgitation will self-progress and increase the inducibility of AF in the long run ^29^.

## Limitations

While this study addresses an important topic with paucity of data on the impact of therapy, it is constrained by its relatively small sample size and single-centre retrospective design. The sample size was limited due to the exclusion of patients with organic mitral valve disease (such as prolapse, flail leaflet, rheumatic disease, or mitral leaflet calcification) and the selection of patients with normal left ventricular morphology and function to minimize the interference caused by ventricular functional regurgitation. Propensity scoring matching could not be applied to compare the SAR and SPR groups due to the limited sample size. Patients with AFMR tend to be older, frailer, have more comorbidities, and have experienced AF for a longer duration. These characteristics may not be as apparent after propensity score matching.

## Conclusions

Since AF is the primary cause of AFMR with AF, rhythm control therapy is the cornerstone treatment for these patients. Thoracoscopic AF procedure is an effective and minimally invasive treatment option for patients with AF and moderate AFMR. Moderate AFMR can be potentially reduced by SR restoration, which promotes cardiac remodeling. For patients with severe AFMR and AF, we recommend concomitant CMP-IV with mitral valvuloplasty. This approach is both safe and effective, increasing the success rate of achieving a resolution of both AF and MR.

## Data Availability

All data are available from the corresponding author on reasonable request.

## Acknowledgements

We would like to thank Mr Yuanzhe Ma, Saie Shen M.D., Ms Huihua Chen and Ms Yu Su for their excellent experimental and clinical support.

